# Relationship between acute SARS-CoV-2 viral clearance with Long COVID Symptoms: a cohort study

**DOI:** 10.1101/2024.07.04.24309953

**Authors:** Carly Herbert, Annukka A.R. Antar, John Broach, Colton Wright, Pamela Stamegna, Katherine Luzuriaga, Nathaniel Hafer, David D McManus, Yukari C Manabe, Apurv Soni

## Abstract

**Introduction:** The relationship between SARS-CoV-2 viral dynamics during acute infection and the development of long COVID is largely unknown.

**Methods:** A total of 7361 asymptomatic community-dwelling people enrolled in the Test Us at Home parent study between October 2021 and February 2022. Participants self-collected anterior nasal swabs for SARS-CoV-2 RT-PCR testing every 24-48 hours for 10-14 days, regardless of symptom or infection status. Participants who had no history of COVID-19 at enrollment and who were subsequently found to have ≥1 positive SARS-CoV-2 RT-PCR test during the parent study were recontacted in August 2023 and asked whether they had experienced long COVID, defined as the development of new symptoms lasting 3 months or longer following SARS-CoV-2 infection. Participant’s cycle threshold values were converted into viral loads, and slopes of viral clearance were modeled using post-nadir viral loads. Using a log binomial model with the modeled slopes as the exposure, we calculated the relative risk of subsequently developing long COVID with 1-2 symptoms, 3-4 symptoms, or 5+ symptoms, adjusting for age, number of symptoms, and SARS-CoV-2 variant. Adjusted relative risk (aRR) of individual long COVID symptoms based on viral clearance was also calculated.

**Results:** 172 participants were eligible for analyses, and 59 (34.3%) reported experiencing long COVID. The risk of long COVID with 3-4 symptoms and 5+ symptoms increased by 2.44 times (aRR: 2.44; 95% CI: 0.88-6.82) and 4.97 times (aRR: 4.97; 95% CI: 1.90-13.0) per viral load slope-unit increase, respectively. Participants who developed long COVID had significantly longer times from peak viral load to viral clearance during acute disease than those who never developed long COVID (8.65 [95% CI: 8.28-9.01] vs. 10.0 [95% CI: 9.25-10.8]). The slope of viral clearance was significantly positively associated with long COVID symptoms of fatigue (aRR: 2.86; 95% CI: 1.22-6.69), brain fog (aRR: 4.94; 95% CI: 2.21-11.0), shortness of breath (aRR: 5.05; 95% CI: 1.24-20.6), and gastrointestinal symptoms (aRR: 5.46; 95% CI: 1.54-19.3).

**Discussion:** We observed that longer time from peak viral load to viral RNA clearance during acute COVID-19 was associated with an increased risk of developing long COVID. Further, slower clearance rates were associated with greater number of symptoms of long COVID. These findings suggest that early viral-host dynamics are mechanistically important in the subsequent development of long COVID.

## Introduction

Long COVID, defined here as the presence of new or persistent symptoms for more than three months after COVID-19 infection that cannot be explained by an alternative diagnosis, is thought to impact between 10%-30% of patients infected with SARS-COV-2.^1–3^ Long COVID varies widely in its manifestation and severity; common symptoms include fatigue, brain fog, and disordered sleep, and affected organ systems include cardiovascular, pulmonary, and neurologic, reviewed in Aiyegbusi et al.^4^ Approximately one-fourth of individuals with long COVID report significant activity limitations, and long COVID has been found to have detrimental effects on quality of life.^5,6^

Currently, the mechanisms leading to long COVID remain unknown. One hypothesis is that persistence of virus or viral antigen may lead to systemic and immunologic dysfunction.^7–9^ Specifically, studies have shown an association between long COVID and delayed clearance of viral RNA during acute COVID-19,^10,11^ persistent SARS-CoV-2 proteins in plasma,^12^ monocytes,^13^ and fecal matter.^14,15^ However, previous studies have been limited by sample size and inconsistent testing frequency.^10,11^

Using a longitudinal cohort of ambulatory participants who were first infected with SARS-CoV-2 between October 2021 and February 2022, we sought to determine if the rate of viral clearance during acute infection is associated with the risk of subsequent development of long COVID. Given that long COVID may comprise two or more mechanistically distinct syndromes, we investigated whether this association exists for some but not all long COVID symptoms. Further, we explored whether specific symptoms displayed during acute SARS-CoV-2 infection were associated with long COVID and the rate of viral clearance.

### Methods

#### Study Population and Data Collection

This study included participants from two longitudinal cohort studies, Test Us at Home and Test Us at Home-Daily, which were funded by the National Institutes of Health’s Rapid Acceleration of Diagnostics (RADx) program. All participants provided written consent to participate, and this study was approved by the WIRB-Copernicus Group (WCG) Institutional Review Board. Participants above the age of two years from across the United States enrolled in these studies remotely between October 2021 and February 2022, and study materials were shipped to participant’s homes. People were eligible to participate if they were asymptomatic on enrollment and owned a smartphone. Communities with a high prevalence of SARS-CoV-2 were targeted for enrollment, to increase the likelihood of capturing transmission and the transition from negative to positive test results. Participants were asked to complete baseline demographic surveys and weekly symptom surveys, as well as collect anterior nasal swabs for SARS-CoV-2 antigen and RT-PCR testing every 48 hours over a 14-day period (Test Us at Home) or every 24 hours over a 10-day period (Test Us at Home-Daily). Detailed information about the parent studies can be found elsewhere.^16^

Participants 18 years of age or older with at least one positive PCR test during the original study period were recontacted in August 2023, 18-22 months after initial SARS-CoV-2 infection. At this follow-up, participants answered a 5-minute survey on persistent symptoms (from a list of 26, see Supplemental Appendix 1) and medical care utilization. Those participants who report no previous SARS-CoV-2 infection on enrollment in the original study were included in this analysis (Figure 1).

**Figure 1:**
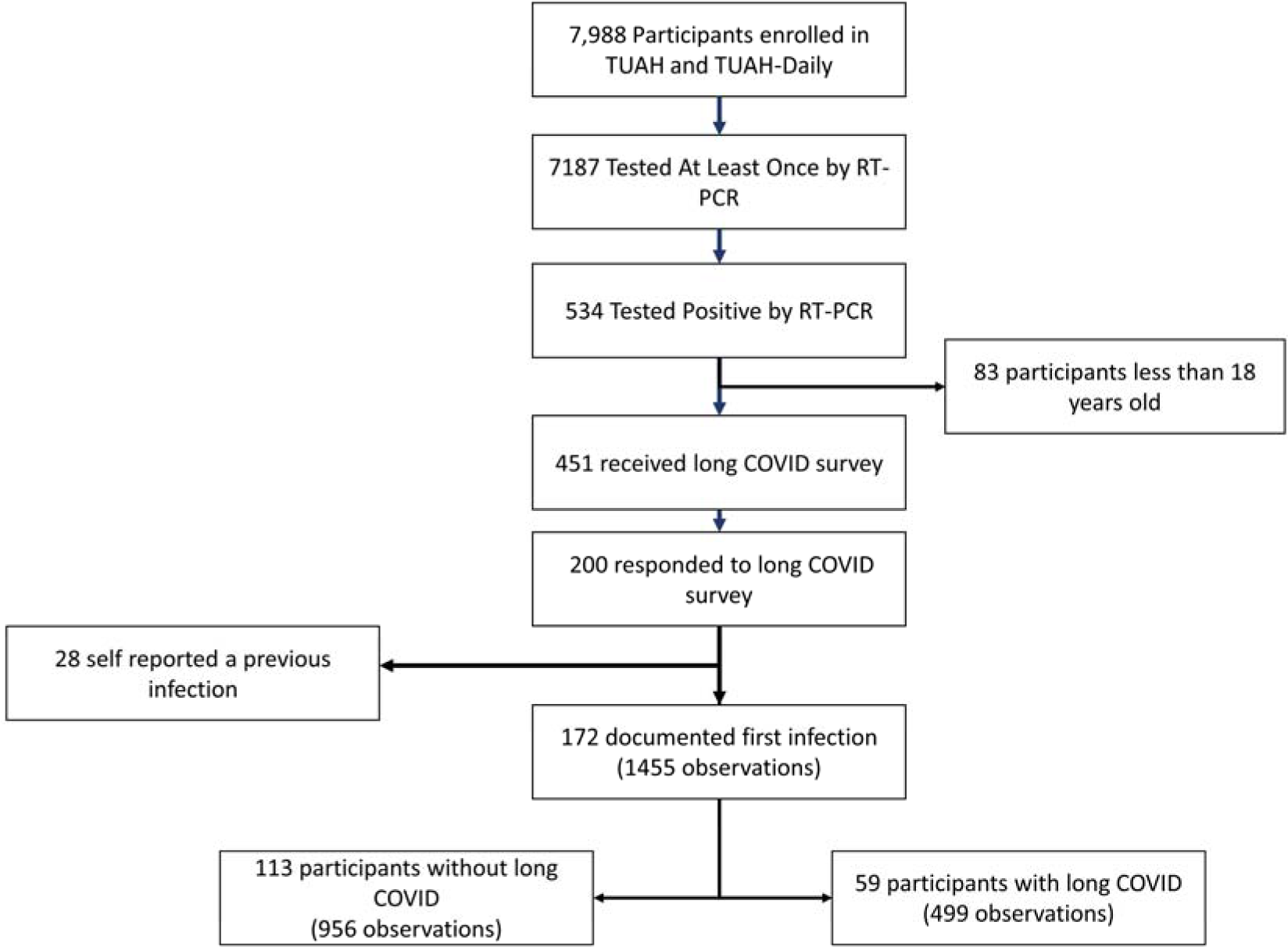
Consort Diagram of Study Cohort. Of the 534 participants who tested positive for SARS-CoV-2 during the parent study, 200 received the long COVID survey and responded. For these analyses, participants were excluded if they were previously infected with SARS-CoV-2; a total of 28 participants self-reported a previous infection and were excluded for this reason. 172 participants with a total of 1455 observations were included in analyses, including 113 participants (65.7%) without long COVID and 59 participants (34.3%) with long COVID.

### Measures

#### SARS-CoV-2 Molecular Testing

Every 24 to 48 hours, participants collected a bilateral anterior nasal swab, which was kept at room temperature and then mailed at room temperature within 24 hours of collection to a central laboratory, Quest Diagnostics, for Roche Cobas RT-PCR testing. Ct values for the E-gene from Roche Cobas 6800 SARS-CoV-2 RT-PCR were used in analyses to quantify viral load. Viral load was calculated as Ln(Viral Load)=((41.44-Ct Value)/1.06), where increasing values represented increased viral load.^17^ Peak viral load was defined as the viral load at the time of the participant’s nadir Ct value.

#### Long COVID Outcomes

In August 2023, participants were asked: “Did you have any symptoms lasting 3 months or longer that you <u>did not have prior to having coronavirus</u> or COVID-19?”.^5^ Participants who answered affirmatively were categorized as having had long COVID and were then asked what symptoms they experienced from among a list of 26 symptoms with a free text box to report more symptoms (Supplemental Appendix 1). We grouped long COVID symptoms by number of symptoms (1-2 long COVID symptoms, 3-4 symptoms, or 5 or more symptoms). All participants were asked about their total number of SARS-CoV-2 infections to date.

#### Survey Data from Original Study

Number of vaccination doses, age, BMI, and comorbidities were self-reported on enrollment. Participants were asked to indicate if they had the following comorbidities: asthma, chronic kidney disease, COPD, cardiovascular disease, cancer, chronic lung disease, depression, diabetes, hypertension, immunocompromising conditions, serious mental health disorders (bipolar or schizophrenia), sickle cell disease, substance use or alcohol use disorders, or other. Participants self-reported acute symptoms (including fever, body aches, fatigue, rash, nausea, abdominal pain, diarrhea, loss of smell, runny nose, cough, headache, or other) immediately prior to each swab collection using the study app. Symptoms were categorized as upper respiratory (loss of smell, runny nose, cough), systemic (fever, body aches, fatigue, headache), or gastrointestinal symptoms (nausea, abdominal pain, diarrhea).

Additionally, symptom reports were used to create a symptom score, which was the sum of symptoms the participant reported on each day. Each participant’s SARS-CoV-2 variant was determined via whole genome sequencing of their positive SARS-CoV-2 sample by amplicon-based next-generation sequencing on extracted RNA.^18^ Non-sequenced positive samples (n=20) were excluded in adjusted analyses.

### Analyses

Demographic characteristics were tabulated and stratified between participants with and without long COVID. To calculate viral clearance, a linear mixed effects model with random slope and random intercept was used to model log viral load values during the period of viral load decline, starting from the peak viral load. To calculate a slope of recovery, participants were required to have two data points: 1) a peak viral load, and 2) one viral load of lesser value occurring after the peak viral load. Log viral load follows a linear pattern over time, which accounts for the exponential phases of viral decline during recovery.^19,20^ Modeled slopes were calculated for each participant (Supplemental Figure 1). Using a log binomial model with the modeled slopes as the exposure, we calculated the relative risk of subsequently developing long COVID with 1-2 symptoms, 3-4 symptoms, or 5+ symptoms. To assess whether the relationship between viral clearance and long COVID was consistent between men and women, we used a log binomial model with an interaction term between slope and biological sex, and nonlinear combinations of predictions were used to quantify the differences by BMI and sex and to calculate the accompanying 95% confidence intervals. We also calculated the relative risk of slope of viral clearance on each long COVID symptom that received more than 5 affirmative responses and displayed these results as a forest plot. We evaluated maximum number of symptoms during acute infection, number of infections, vaccination status, age, comorbidities, BMI, sex, and SARS-CoV-2 variant as confounders, and included variables based on a 10% change-in-estimate criterion.^21^ Maximum symptoms, age, and variant were determined to be confounders using this criterion and included as confounders in the adjusted models. To facilitate interpretation, we also displayed slopes in terms of days to clearance, assuming intercepts (i.e., maximum viral load) at the 25^th^, 50^th^, and 75^th^ percentile. Estimates and 95% confidence intervals were generated through the bootstrap methodology to incorporate measures of uncertainty. Clearance was defined as a Ct value ≥40 which is the limit of detection (i.e., log viral load of 3.90).

Lastly, we analyzed participant-reported symptoms recorded during acute infection and calculated relative risk of subsequent development of long COVID for each individual acute symptom, acute symptom category (upper respiratory, gastrointestinal, or systemic), and cumulative number of acute symptoms. Confounders were similarly determined using a 10% change-in-estimate criterion, and models were adjusted for age and variant. We conducted sensitivity analyses analyzing participants who reported just one total SARS-CoV-2 infection as of August 2023. All analyses were conducted using STATA 17.0.

## Results

### Participant Characteristics

Of the 7988 participants enrolled in parent studies Test Us at Home and Test Us at Home Daily, 451 participants tested positive for SARS-CoV-2 by RT-PCR during the parent study and were 18 years or older, and thus eligible for the long COVID survey in August 2023. In total, 200 participants (44.3%) responded to the long COVID survey. We limited analyses to the 172 participants whose first known infection with SARS-CoV-2 occurred during the parent study period (**Figure 1**). Among 172 respondents, 59 (34.3%) reported having had long COVID, here defined as the presence of symptoms for three months or longer following COVID-19.

In total, 128 of the 172 (74.4%) of participants were women, and the median age was 37 (IQR, 32-44). The majority of participants (62.2%) had no comorbidities, while 11.0% of participants had 2+ comorbidities (**Table 1**). Among the 59 participants with long COVID, 8 (13.6%) visited the emergency room during their SARS-CoV-2 infection, compared to just 1 participant (0.88%) without long COVID. Only one participant was hospitalized during their acute SARS-CoV-2 infection. As of August 2023, 71.7% of participants who never had long COVID reported having just one SARS-CoV-2 infection, while 40.7% of those with long COVID reported having just one SARS-CoV-2 infection. In our sample, 128 participants (74.4%) had their peak viral load and a prior lower viral load captured during the study period.

**Table 1:** Demographics of Cohort by Long COVID Status.

|  |  | No Long COVID<br>N=113 | Long COVID<br>N=59 | P-value |
| --- | --- | --- | --- | --- |
| Sex | Female | 82 (72.6%) | 46 (78.0%) | 0.44 |
|  | Male | 31 (27.4%) | 13 (22.0%) |  |
| Acute COVID-19 Severity | No symptoms | 6 (5.3%) | 0 (0.0%) | <0.001 |
|  | Managed symptoms at home | 104 (92.0%) | 51 (86.4%) |  |
|  | Visited the emergency department | 1 (0.88%) | 8 (13.6%) |  |
|  | Hospitalized | 1 (0.88%) | 0 (0.0%) |  |
|  | I don't remember | 1 (0.88%) | 0 (0.0%) |  |
| Vaccination Doses Prior to First SARS-CoV-2 infection | Unvaccinated | 23 (20.4%) | 7 (11.9%) | 0.14 |
|  | 1 dose | 4 (3.5%) | 1 (1.7%) |  |
|  | 2 doses | 19 (16.8%) | 18 (30.5%) |  |
|  | 3+ doses | 67 (59.3%) | 33 (55.9%) |  |
| BMI Categories | BMI <25 | 47 (41.6%) | 17 (28.8%) | 0.24 |
|  | BMI 25-30 | 31 (27.4%) | 18 (30.5%) |  |
|  | BMI >30 | 35 (31.0%) | 24 (40.7%) |  |
| Number of COVID-19 infections | 1 | 81 (71.7%) | 24 (40.7%) | <0.001 |
|  | 2 | 22 (19.5%) | 28 (47.5%) |  |
|  | 3 | 7 (6.2%) | 7 (11.9%) |  |
|  | 4 | 3 (2.7%) | 0 (0.0%) |  |
| Age Category | 18-44 years | 88 (77.9%) | 38 (64.4%) | 0.022 |
|  | 45-64 years | 17 (15.0%) | 18 (30.5%) |  |
|  | 65+ years | 4 (3.5%) | 0 (0.0%) |  |
|  | Missing | 4 (3.5%) | 3 (5.1%) |  |
| Comorbidities<sup>a</sup> | None | 73 (64.6%) | 34 (57.6%) | 0.66 |
|  | 1 comorbidity | 28 (24.8%) | 18 (30.5%) |  |
|  | 2+ comorbidities | 12 (10.6%) | 7 (11.9%) |  |
| SARS-CoV-2 Variant | Omicron | 92 (84.4%) | 44 (78.6%) | 0.35 |

Data are presented as n (%).
<sup>a</sup>Comorbidities included: asthma, chronic kidney disease, COPD, cardiovascular disease, cancer, chronic lung disease, depression, diabetes, hypertension, immunocompromising conditions, serious mental health disorders (bipolar or schizophrenia), sickle cell disease, substance use or alcohol use disorders.

### Association between long COVID and Slope of Viral Clearance

The slope of viral clearance was higher (i.e., flatter) among participants with 5+ long COVID symptoms compared to those with 3-4 long COVID symptoms, 1-2 long COVID symptoms, or those without long COVID (**Figure 2a**, **Figure 2b**). Slope of viral clearance had no association with risk of long COVID with only 1-2 symptoms (**Table 2**). However, the risk of long COVID with 3-4 symptoms was 2.44 times higher (Adjusted RR (aRR): 2.44; 95% CI: 0.88-6.82) in models adjusted for age, maximum acute symptoms, and SARS-CoV-2 variant per slope-unit increase (**Table 2**). This association was strongest among participants with 5+ long COVID symptoms, where a slope-unit increase was associated with nearly five-times higher risk of long COVID with 5+ symptoms (aRR: 4.97; 95% CI: 1.90-12.98). The interaction term for biological sex was statistically significant (p=0.023), and we observed that the relationship between slope and long COVID was not consistent between men and women. While the probability of long COVID increased as slope decreased among females, the same relationship was not observed among men (**Figure 3**).

**Figure 2:**
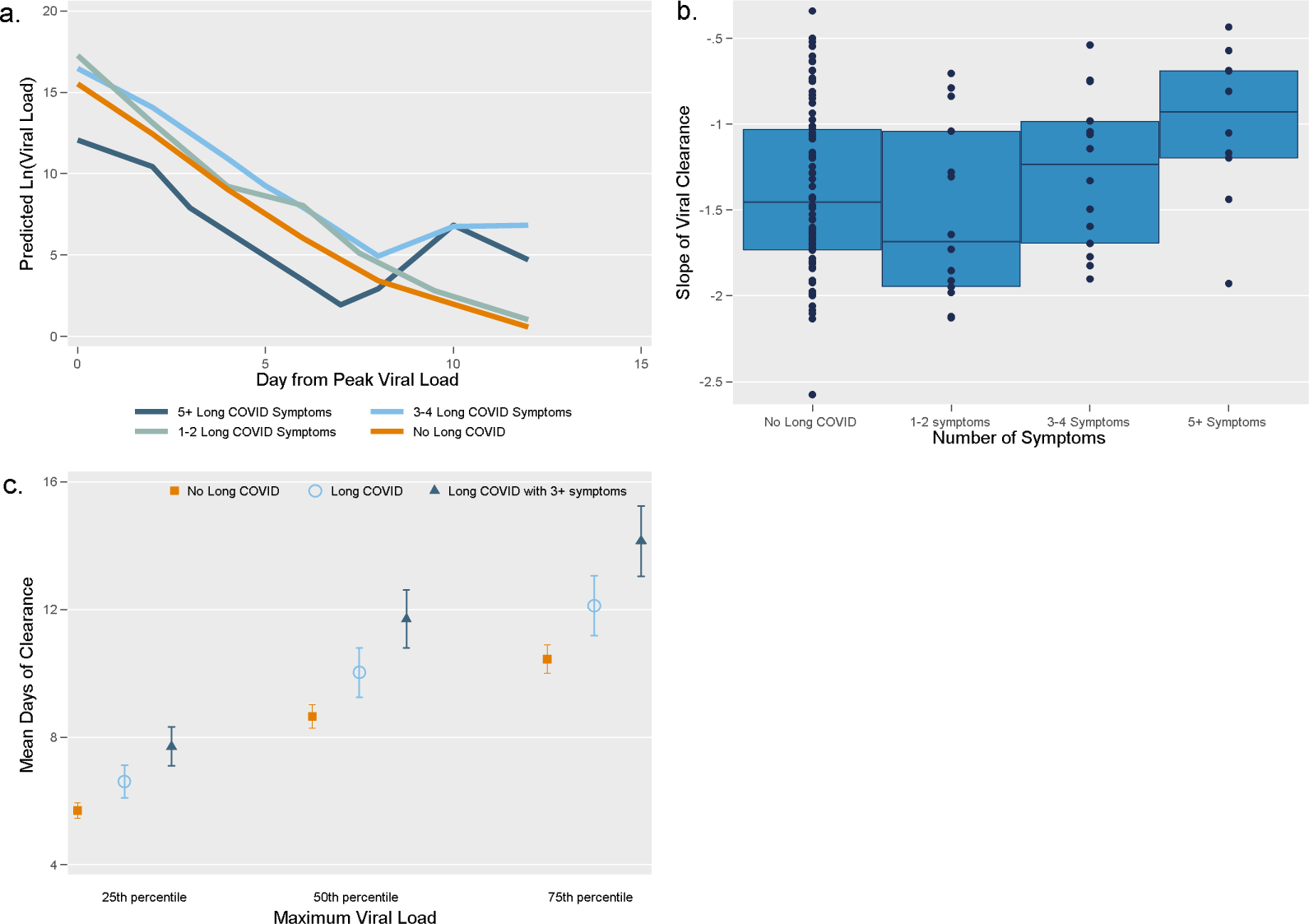
Predicted Viral Load, Slope, and Time to Clearance by Number of Long COVID Symptoms. **Figure 2a shows mean predicted viral load each day starting from the day of peak viral load by number of long COVID symptoms.** Dark blue signifies individuals with 5+ long COVID symptoms, light blow shows individuals with 3-4 long COVID symptoms, grey indicates individuals with 1-2 long COVID symptoms, and the orange line includes individuals with no long COVID. Figure 2b displays the slope of viral clearance among participants with no long COVID, 1-2 long COVID symptoms, 3-4 long COVID symptoms, and 5+ long COVID symptoms. Higher slopes (closer to 0) are “flatter” than lower slopes (more negative). The blue boxes represent the interquartile range, and the navy line within the blue box displays the median slope for each group. Navy dots show the slope for each participant. Figure 2c shows predicted mean number of days from peak viral load to clearance calculating using the slopes of viral clearance, assuming clearance at a Ct value=40 (limit of detection). Days from peak viral load to clearance is a function of peak viral load and speed (slope) of clearance, so these estimates were calculated using maximum viral loads at the 25^th^ percentile, 50^th^ percentile, and 75^th^ percentile. Participants with long COVID with 3+ symptoms are shown in dark blue triangles, those with long COVID (with any number of symptoms) are shown in light blue circles, and participants without long COVID are in orange squares. Whiskers represent 95% confidence intervals.

**Figure 3:**
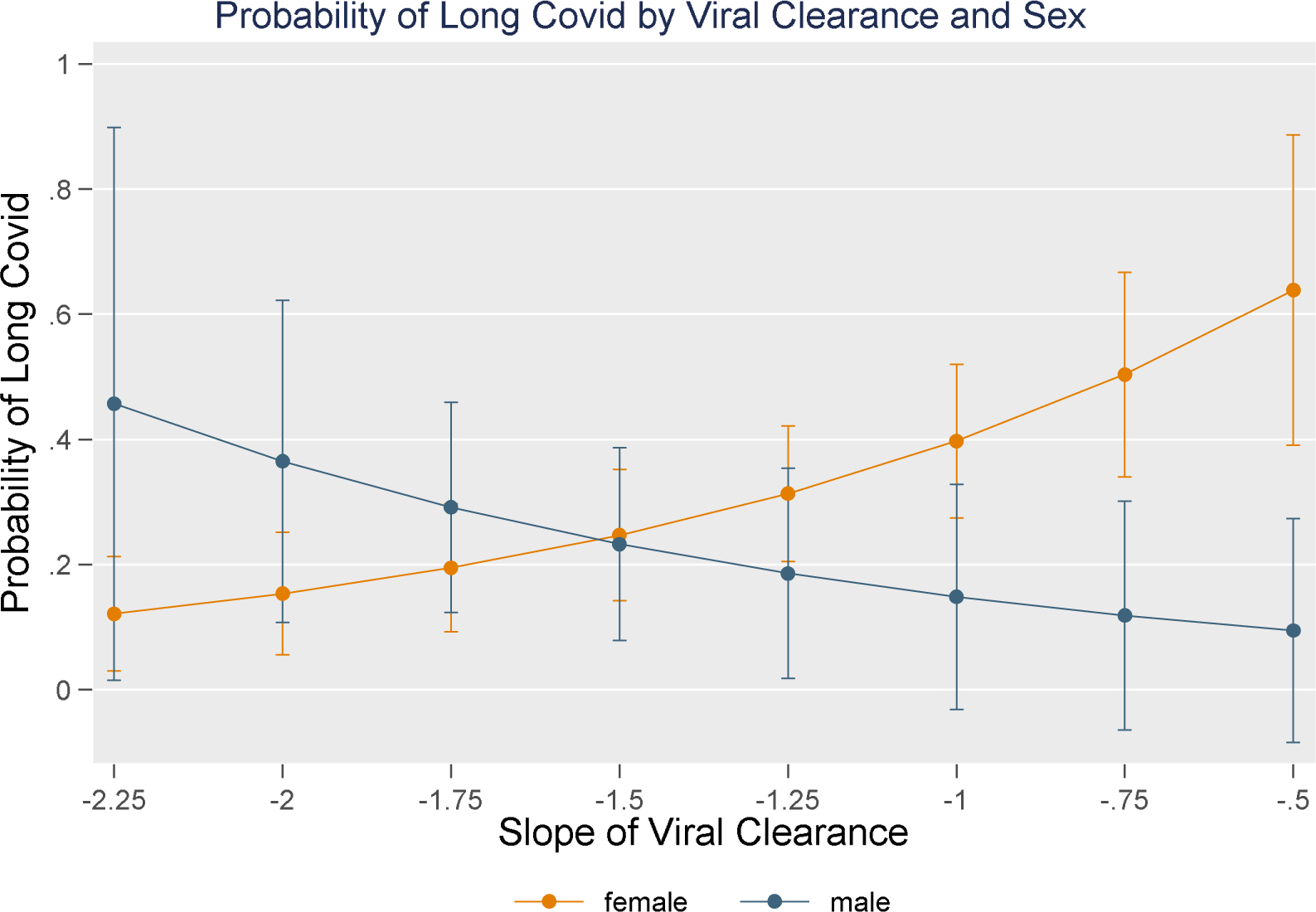
Probability of Long COVID by Viral Clearance and Sex. Females are displayed in orange and males are displayed in blue. Whiskers show 95% confidence intervals. As slope increases, probability of long COVID increases among females. However, there is no significant increase in probability of long COVID as slope increases among males.

**Table 2:** Association of Slope of Viral Clearance with Number of Long COVID Symptoms. Adjusted estimates are adjusted for maximum symptoms, age, and SARS-CoV-2 variant (Delta or Omicron). A unit-increase in slope was associated with a 4.97 times higher risk of having 5+ long COVID symptoms, adjusted for maximum symptoms during acute SARS-CoV-2 infection, age, and variant.

| Number of Long COVID Symptoms | Slope of Viral Clearance:<br>$\beta$ (95% CI) | |
| --- | --- | --- |
|  | Unadjusted Relative Risk | Adjusted* Relative Risk |
| No Long Covid<br>(reference group) | 1 | 1 |
| 1-2 symptoms | 0.59 (0.21-1.69) | 0.65 (0.22-1.97) |
| 3-4 symptoms | 1.49 (0.55-4.01) | 2.44 (0.88-6.82) |
| 5+ symptoms | 4.39 (1.27-15.23) | 4.97 (1.90-12.98) |

For participants who never had long COVID, the time from maximum viral load to viral clearance during the first SARS-CoV-2 infection was 8.65 days (95% CI: 8.28-9.01), assuming a nadir Ct value of 24.2, which was the median among this population (**Figure 2c, Supplemental Table 1**). Participants with long COVID had significantly longer times to viral clearance in their first SARS-CoV-2 infection than those without long COVID (No long COVID: 8.65 (95% CI: 8.28-9.01); Long COVID: 10.0 (95% CI: 9.25-10.8)). For participants with more than three long COVID symptoms, time to clearance was highest, at 11.7 days (95% CI: 10.8-12.6). For participants with peak viral loads in the 25^th^ percentile (nadir Ct value=28.6), time to clearance was lower, and ranged from 5.70 days (95% CI: 5.46-5.94) among those without long COVID to 7.71 days (95% CI: 7.11-8.32) among those with more than three long COVID symptoms. For participants with peak viral loads in the 75^th^ percentile (nadir Ct value=21.4), time to clearance was higher, and ranged from 10.5 days (95% CI: 10.0-10.9) among those without long COVID to 14.2 days (95% CI: 13.0-15.3) among those with three or more long COVID symptoms.

### Relationship between Slope of Viral Clearance and Specific Long COVID Symptoms

Slope of viral clearance was significantly positively associated with long COVID symptoms of fatigue (aRR: 2.86; 95% CI: 1.22-6.69), brain fog (aRR: 4.94; 95% CI: 2.21-11.0), shortness of breath (aRR: 5.05; 95% CI: 1.24-20.6), and gastrointestinal symptoms (aRR: 5.46; 95% CI: 1.54-19.3) (**Figure 4**). In other words, for each unit increase in slope, the risk of long COVID with fatigue increased by 2.86 times. We did not observe a significant association between slope of viral clearance and long COVID symptoms of cough, change in taste or smell, mental health symptoms, or hair loss.

**Figure 4:**
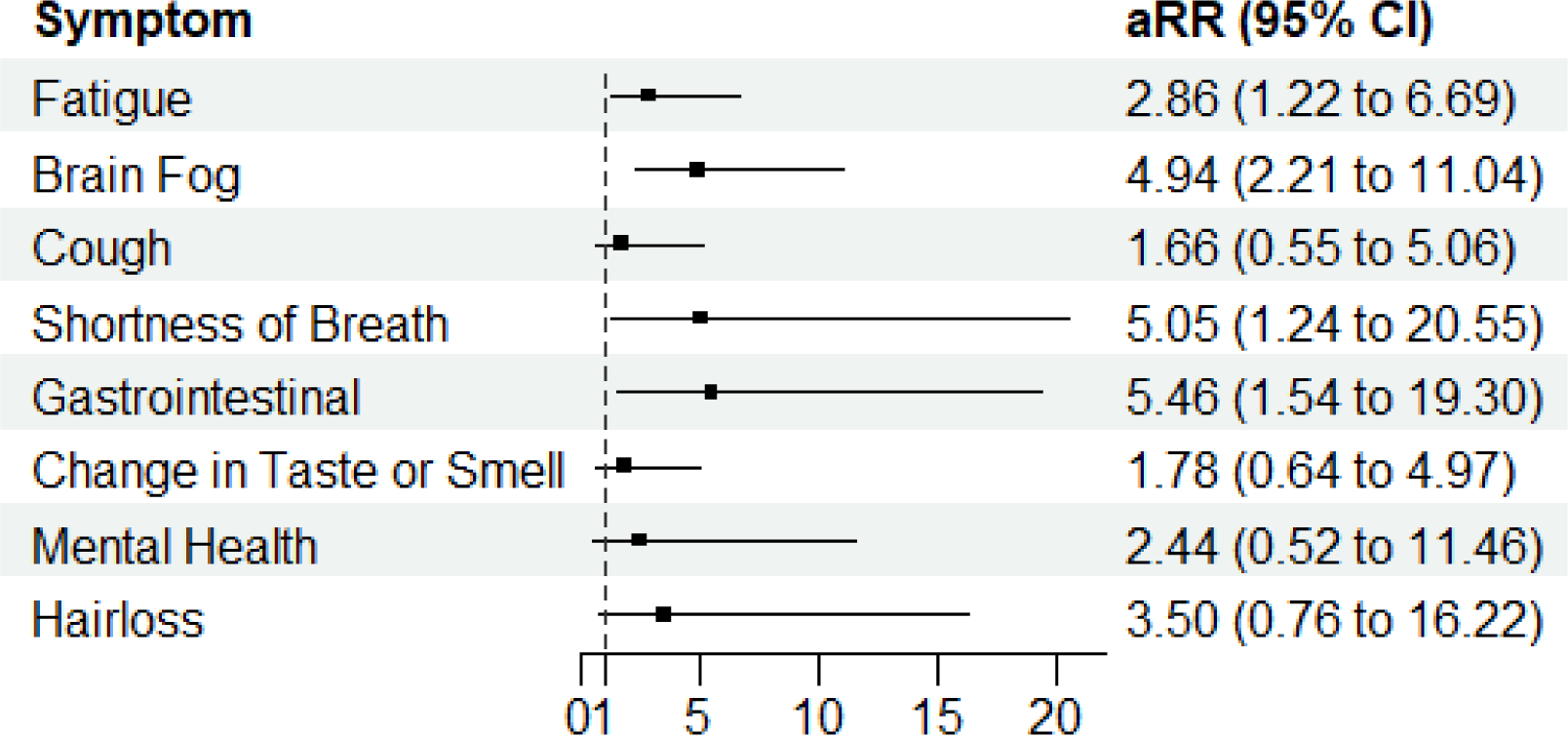
Association of Specific Long COVID Symptoms with Slope of Viral Clearance. Dotted black line shows a relative risk of 1. Black dots show the relative risk of each long COVID symptom from slope of viral clearance, adjusted for maximum number of symptoms during acute infection, age, and SARS-CoV-2 variant (Delta or Omicron). aRR: Adjusted Relative risk

### Acute COVID-19 Symptoms and Risk of long COVID

Each additional symptom reported during the acute COVID-19 infection period was associated with a 21% (95% CI: 1.03-1.43) and 24% (95% CI: 1.11-1.39) increased risk of long COVID with 3-4 symptoms and 5+ symptoms, respectively (**Table 3**). No acute symptoms were associated with increased risk of long COVID with 1-2 symptoms. We observed that the acute symptoms of abdominal pain (aRR: 5.41; 95% CI: 2.44-12.0), nausea (aRR: 3.01; 95% CI: 1.31-6.89), and body aches (aRR: 2.58; 95% CI: 1.26-5.30) during SARS-CoV-2 infection were the three individual symptoms most strongly associated with long COVID (Figure 5). When looking at symptom groupings, upper respiratory symptoms during acute infection, including loss of smell, runny nose, and cough, were not associated with an increased risk of long COVID, regardless of the number of symptoms. Gastrointestinal symptoms were associated with 3.91 times the risk of long COVID with 5+ symptoms after adjusting for age, BMI, and variant (aRR: 3.91, 95% CI: 1.72-8.92); however, gastrointestinal symptoms were not significantly associated with an increased risk of long COVID with 3-4 symptoms. Furthermore, systemic symptoms were associated with 4.16 times (95% CI: 1.63-10.6) the risk of long COVID with 3-4 symptoms and 5.52 times (95% CI: 2.02-15.0) the risk of long COVID with 5+ symptoms. These results were consistent when restricting to patients who reported just one SARS-CoV-2 infection as of August 2023 (**Supplemental Table 2**).

**Figure 5:**
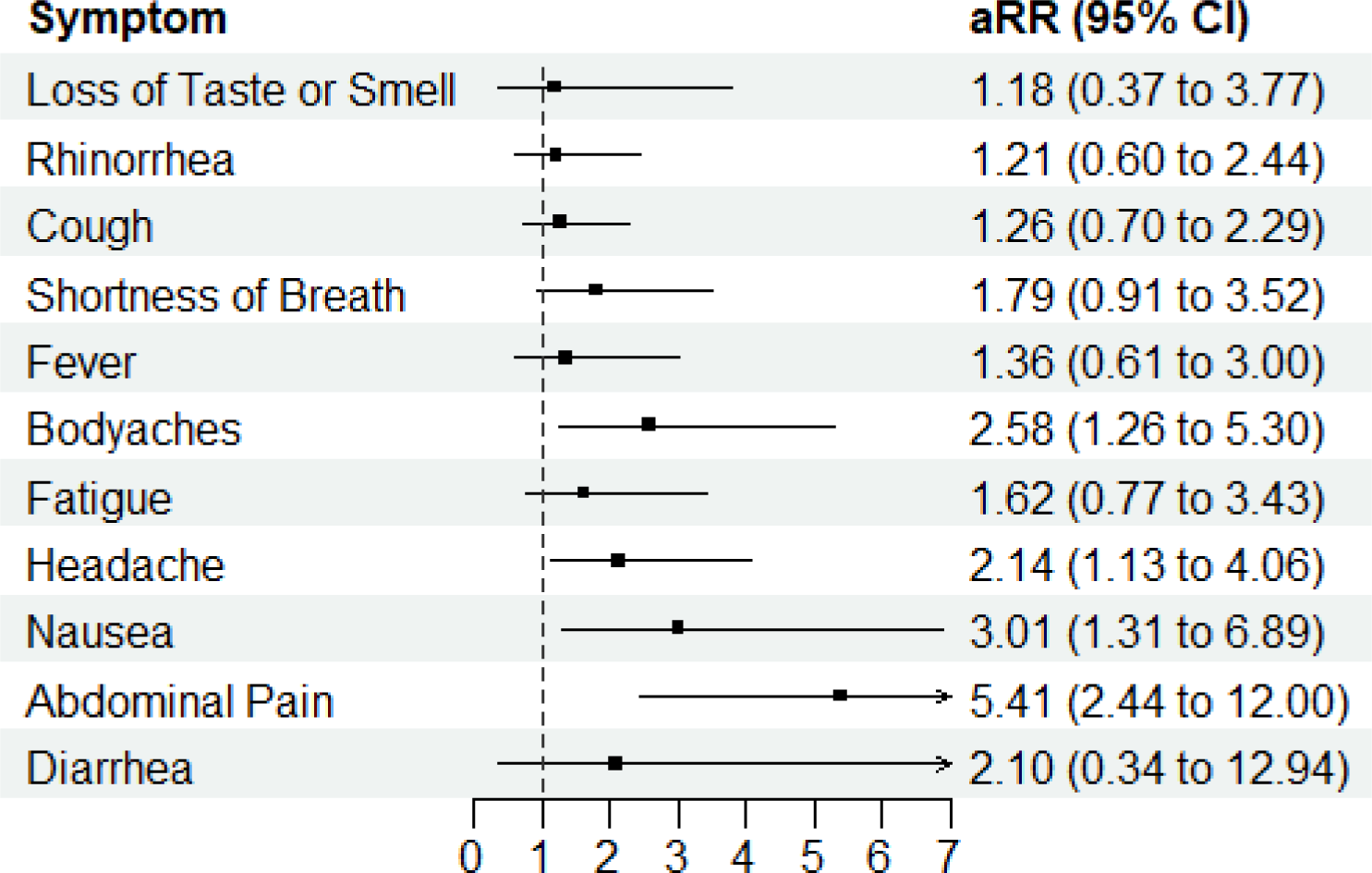
Association Between Acute SARS-CoV-2 Symptoms and Risk of Long COVID. Dotted black line shows a relative risk of 1. Black dots show the adjusted relative risk of long COVID from acute SARS-CoV-2 symptoms, adjusted for age, BMI, and SARS-CoV-2 variant (Delta or Omicron). aRR: Adjusted Relative risk

**Table 3:**
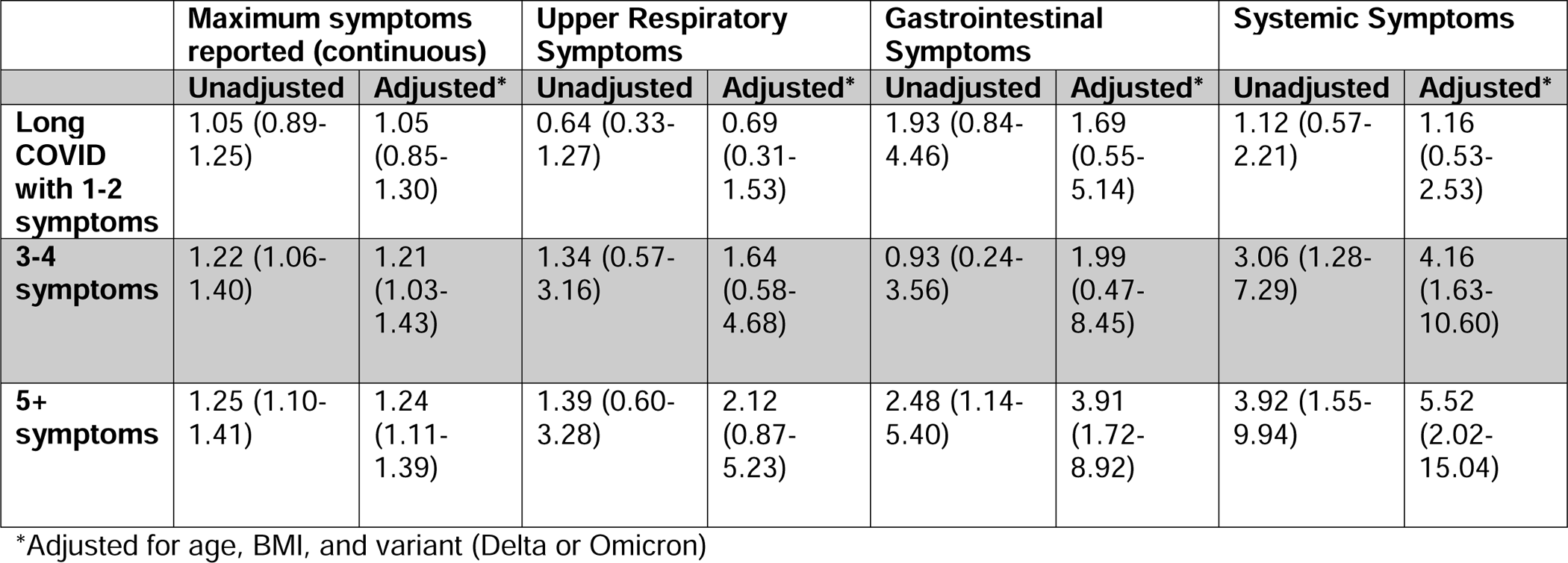
Association between Number of Acute COVID-19 Symptoms and Risk of Long COVID.

## Discussion

Using longitudinal data collected from participants during their acute SARS-CoV-2 infections, we report that longer time to clearance during acute COVID-19 is associated with an increased risk of long COVID, and this relationship is stronger with increasing numbers of long COVID symptoms. This study is unique in its ability to pair acute infection dynamics with long term outcomes of COVID-19 infection; we offer a prospective, longitudinal examination of infection trajectories and their relationship with long COVID. Our large sample size includes participants from across the United States and includes infections with both the Omicron and Delta SARS-CoV-2 variants. Further, the frequent sample collection during the acute SARS-CoV-2 infection period allowed us to model viral clearance with high granularity.

### Delayed SARS-CoV-2 Viral Clearance and Long COVID

Most studies of clinical viral dynamics have occurred in hospital settings; however, nearly 81% of SARS-CoV-2 infections are mild to moderate and self-resolving.^22^ Additionally, the majority of long COVID patients were not hospitalized for COVID-19.^23^ Therefore, our study of adults with mild to moderate infections offers an important lens into the longitudinal course of SARS-CoV-2 infections occurring outside the clinical setting. Among ambulatory outpatients with mild to moderate SARS-CoV-2 infections, Antar et al. also found that delayed time to clearance (defined as viral clearance >28 days after symptom onset) was associated with long COVID with brain fog and muscle pain.^10^ However, they did not find a statistically significant association between viral clearance and other long COVID symptoms, likely due to sample size and less frequent sampling. Our larger sample size, more frequent sampling, and inclusion of participants who went from SARS-CoV-2 negative to positive, allowed us to use a continuous measure to show that time to clearance was longer among those with long COVID than those without long COVID, rather than categorizing clearance at 28 days. Similarly, we found a strong association between viral clearance and brain fog, as well as GI symptoms and shortness of breath. Another study of participants with mild to moderate SARS-CoV-2 infections also found an association between long COVID and higher peak viral loads, as well as duration of viral shedding.^11^ This study included primarily participants infected with non-Omicron variants, and 65% of their participants were unvaccinated. Therefore, it is notable that we found that time to viral clearance had an association with long COVID in our population too, which included majority Omicron infections and vaccinated participants. Additionally, our study had >50% more participants, and we were uniquely able to document the transition from negative to positive, while Lu et al. enrolled participants within 5 days of their first positive test.

Our results add to evidence showing an association between delayed viral clearance and long COVID.^10,11^ Delayed viral clearance may be related to long COVID through several mechanisms. First, delayed immune response, specifically secretory IgA, to SARS-CoV-2 may cause delayed viral clearance.^24^ This may occur due to underlying immune dysfunction caused by autoimmune or comorbid conditions, which themselves may be associated with long COVID.^25,26^ Furthermore, our finding is consistent with the finding that vaccination provides partial protection from long COVID.^25^ In this case, vaccination may enable an infected individual to mount a faster or more effective immune response to clear viral RNA, and in turn this faster clearance is associated with decreased risk for long COVID.^27–30^ Additionally, delayed viral clearance may in itself *cause* immune dysfunction through chronic immune activation, including lymphocyte overstimulation, leading to inflammatory responses and potential cross-reactive immune responses.^31–33^ Whether the immune dysfunction underlies or causes the delayed viral clearance is still unknown; however, this heightened inflammatory state may possibly delay healing from the acute infection, trigger autoimmunity, or cause organ damage, all resulting in long-term symptoms. Therefore, the speed and functionality of the immune response responsible for clearing virus from the upper respiratory tract may either directly impact long COVID risk or be associated with other dysfunction or damage related to long COVID, and result in serious implications for long-term symptom resolution.^31,32^

We also observed that the association between viral clearance and long COVID differed between men and women. It appeared the relationship between viral clearance and long COVID was stronger among women, compared to men. It is well documented that men comprised more COVID-19-related deaths and hospitalizations, compared to women, and evidence has shown higher viral loads among men compared to women.^34,35^ However, women are more likely than men to report long COVID.^1,36^ Men and women have been observed to have different responses to viral infection, with men having less robust innate immune responses than women.^37,38^ This may indicate that the immune dysfunction leading to long COVID is mediated by sex-specific factors; however, this observation may also be due to the limited number of men in our study cohort. Therefore, further sex-specific analyses are necessary to further evaluate this finding.

### Association between acute symptoms and Long COVID

We found that certain symptoms during acute COVID-19 infection were associated with a higher risk of long COVID than others.^39,40^ Abdominal pain during acute COVID-19 infection was the single symptom most significantly associated with future risk of long COVID. Gastrointestinal symptoms are reported in 15-50% of acute COVID-19 infections, and SARS-CoV-2 RNA, antigen, and virions have all been identified in the GI tract of COVID-19 patients.^41–43^ Further, previous studies have indicated that SARS-CoV-2 clearance in respiratory tissues may be more rapid than clearance within gastrointestinal tissues.^14^ Fecal shedding of RNA has been correlated with a variety of GI symptoms, including abdominal pain. It may be that abdominal pain is a crude indicator of viral dissemination outside the upper respiratory tract, and viral dissemination is associated with long COVID. However, viral RNA in the GI tract has been found in patients with and without long COVID.^14^ The GI tract is highly immunoreactive, and it is hypothesized that prolonged exposure to SARS-CoV-2 in the GI tract may have immunological implications and contribute to many common long COVID symptoms.

### Limitations and strengths

This study has several limitations. The parent study period was 14 days; therefore, duration of infection was calculated through modeled estimates, rather than observed clearance. While we classified long COVID by number of symptoms reported, we did not consider the severity of each symptom. We were also limited in our ability to perform further subgroup analyses on sex and long COVID symptoms due to sample size. Previous infections were self-reported; therefore, it is possibly participants were unknowingly infected with SARS-CoV-2 prior to the infection documented during the study period. There is also the potential for recall bias for long COVID symptoms. Further, the FDA granted an EUA for Paxlovid for the treatment of individuals with mild to moderate COVID-19 at risk of developing severe COVID-19 on December 22, 2021, which occurred in the middle of our study. However, outside of long-term care settings, use of Paxlovid in January and February 2022 was very low in the United States, with less than 22 prescriptions and less than 40 prescriptions per 100,000 people <65 years old and >65 years old, respectively.^44^ Lastly, as of December 2022, more than 50% of COVID-19 cases in the United States were reinfections, and individual’s cumulative exposure to the virus continues to increase with the mutation and spread of the Omicron variant.^45^ Understanding the viral dynamics from an individual’s first COVID-19 infection can form a baseline to enhance our understanding of the relationship between subsequent infections on the risk of long COVID.^46^ However, future studies are needed to explore the impact of reinfection on viral clearance and the connection with long COVID.

### Conclusion

We observed that slower rates of viral RNA clearances during acute COVID-19 were associated with an increased risk of developing long COVID. Further, slower clearance rates were associated with a greater number of long COVID symptoms. These findings suggest that early viral-host dynamics are mechanistically important in the subsequent development of long COVID.

## Data Availability

All data produced in the present study are available upon reasonable request to the authors.

https://radxdatahub.nih.gov/

## Patient Consent Statement

All participants provided written consent for this study. This study was approved by the WIRB-Copernicus Group (WCG) Institutional Review Board (20214875).

## Author Contributions

YCM, AA, AS, CH conceptualized the research question. CH, AS, YCM, AA contributed to design of methodology. AS, CH, CW, JB, PS contributed to data collection and curation. CH conducted formal analysis and wrote the original draft. KL, DDM, NH, YCM, AA provided supervision. CW and PS provided administrative support. DM, JB, AS, NH, KL contributed to funding acquisition. All authors contributed to reviewing and editing the manuscript.

## Acknowledgment

We are grateful to our study participants and to our collaborators from the National Institute of Health (NIBIB and NHLBI) who provided scientific input into the design of this study and interpretation of our results but could not formally join as co-authors due to institutional policies and to the Food and Drug Administration (Office of In Vitro Diagnostics and Radiological Health) for their involvement in the primary TUAH study. We received meaningful contributions from Drs. Bruce Tromberg, Jill Heemskerk, Felicia Qashu, Dennis Buxton, Erin Iturriaga, Jue Chen, Andrew Weitz, and Krishna Juluru. We are thankful to county health departments across the country who helped with recruitment for this siteless study by spreading the word in their networks.

