## Supplemental Tables and Figures for "Relationship between acute SARS-CoV-2 viral clearance with Long COVID Symptoms: a cohort study"

***Supplemental Material***

### **Supplemental Figure 1: Actual and Modeled Cycle Threshold Values by Day Post-Cycle threshold Nadir**

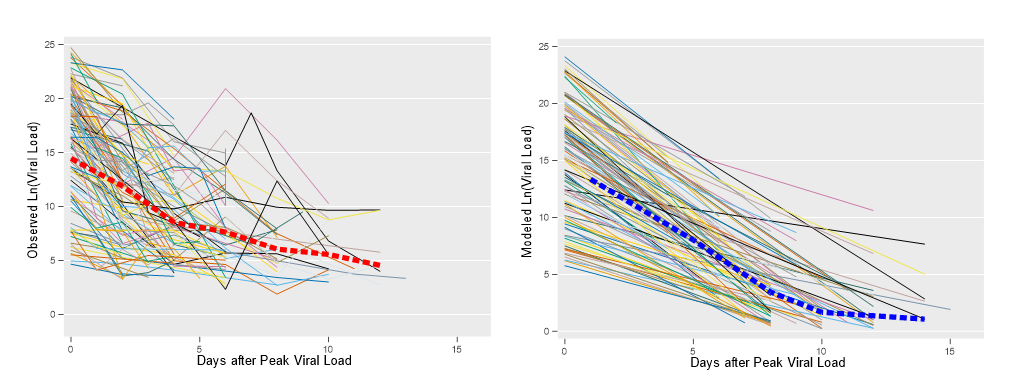

**Actual and Modeled Cycle Threshold Values starting on Day of Peak Viral Load.** The image to the left shows the observed ln(viral load) starting on the day of peak viral load. Each colored line represents one participant, and the thick, red, dashed line represents the population mean ln(viral load). The image to the right shows the modeled ln(viral load) based on the mixed- effects model with random intercept and slope for each participant. The colored lines show the modeled ln(viral load) for each participant, and the thick, blue, dashed line shows the modeled population mean viral load per day.

### **Supplemental Table 1: Days From Viral Peak to Clearance by Number of Long COVID Symptoms**

|  | Days from Viral Peak to Clearance | | |
| --- | --- | --- | --- |
|  | 25^th^ percentile peak viral load | 50^th^ percentile peak viral load | 75^th^ percentile peak viral load |
| No Long COVID | 5.70 (5.46-5.94) | 8.65 (8.28-9.01) | 10.45 (10.01-10.89) |
| Any Long COVID | 6.61 (6.10-7.12) | 10.03 (9.25-10.80) | 12.12 (11.19-13.05) |
| Long COVID with 3+ symptoms | 7.71 (7.11-8.32) | 11.70 (10.79-12.62) | 14.15 (13.04-15.25) |

**Days From Viral Peak to Clearance by Number of Long COVID Symptoms.** Clearance was defined as a Ct value ≥40 (i.e., log viral load of 3.90), and time to clearance was calculated using individual modeled viral slopes. Estimates and 95% confidence intervals were generated through the bootstrap methodology to incorporate measures of uncertainty.

### **Supplemental Table 2: Association of Acute Symptoms with Risk of Long COVID among Individuals with 1 Known COVID-19 Infection**

|  | Relative Risk of Long COVID | | | |
| --- | --- | --- | --- | --- |
|  | β | 95% CI | Adjusted | 95% CI |
| Maximum symptoms reported (continuous) | 1.15 | 1.04-1.28 | 1.21 | 1.06-1.39 |
| URI symptoms | 0.74 | 0.42-1.31 | 0.87 | 0.46-1.66 |
| GI symptoms | 2.12 | 1.13-3.99 | 2.90 | 1.35-6.24 |
| Systemic symptoms | 1.78 | 0.99-3.19 | 2.45 | 1.21-4.95 |

**Association of Acute Symptoms with Risk of Long COVID among Individuals with 1 Known COVID-19 Infection.** Analysis restricted to just those with one COVID infection. Individuals with reinfections have been excluded. Adjusted estimates are adjusted for age, BMI, and variant (Delta or Omicron). URI: upper respiratory symptoms; GI: gastrointestinal.
